# Remission of type 2 diabetes can be achieved in short and long-lasting type 2 diabetes: results of a randomized controlled trial in individuals with overweight and obesity

**DOI:** 10.1101/2025.06.10.25329355

**Authors:** Bettina Schuppelius, Elena Lalama, Jiudan Zhang, Kilian Rüther, Marta Csanalosi, Stefan Kabisch, Anette Christ, Eicke Latz, Olga Pivovarova-Ramich, Knut Mai, Andreas F. H. Pfeiffer

## Abstract

**Background and Aim:** A growing body of evidence shows that upon extensive weight loss returning to non-diabetic glucose control is possible in people with recent onset type 2 diabetes (T2D). However, the impact of diabetes duration and different intervention strategies on remission is not clear. Thus, we investigated the remission of T2D in response to three months of two very low-calorie diets (VLCDs) with different macronutrient profiles in individuals with short and long diabetes duration.

**Methods:** Participants with a BMI >27 kg/m^2^ and T2D duration of ≤4 years or ≥8 years were studied before and after following a VLCD strategy (600-800 kcal/day) for three months including discontinuation of antidiabetic medication. Individuals were stratified by diabetes duration and randomly assigned to one of two VLCDs with slightly different macronutrient composition. Phenotyping included mixed meal tolerance test (MMTT), metabolic characterization and assessment of body composition.

**Results:** Fifty-two (30 women, 22 men) participants were enrolled between September 2020 and November 2022 into the trial and 47 participants completed the intervention. Remission of T2D, defined as plasma fasting glucose levels <126 mg/dl, was achieved in 34 participants (72%). Despite similar weight loss of subjects with a diabetes duration ≤4 years and ≥8 years (-15.2 ± 5.8 kg vs. -13.9 ± 4.8 kg; p = 0.473), subjects with diabetes duration ≥8 years had a 32% lower remission rate (82% vs. 50%; p = 0.027). Remission rates also differed between the two formula diets and were found to be higher with the high-fiber, high-protein, and low-carb, low-fat formula diet (91% vs. 56%; p = 0.008). In addition, individuals that achieved a remission had significantly lower fasting plasma glucose and higher C-peptide levels at baseline.

**Conclusions:** Our findings show that significant body weight loss through VLCDs can induce T2D remission in nearly three-quarters of participants. Fasting plasma glucose and C-peptide levels, diabetes duration, and used macronutrient profile emerged as important factors for the achievement of diabetes remission, although a considerable remission is still possible after a long duration of T2D.

## Introduction

Over the past 15 years, a growing body of evidence showed that a negative energy balance and concomitant weight loss can reverse T2D in lean individuals [1] as well as individuals with overweight or obesity [2–4]. However, this T2D remission cannot be achieved in all individuals. Individual patient characteristics or the impact of specific interventional strategies are not well analyzed so far.

Currently, no particular macronutrient composition or hypocaloric diet type was shown to be superior to reduce body weight in individuals with T2D [3–5]. To induce a remission of T2D, very low-calorie diets (VLCD) using total diet replacement strategies were shown to be very effective [3]. Besides lowering plasma glucose levels into the non-diabetic range, the restricted energy intake led to an average weight loss of 15 kg, reduced hepatic and pancreatic fat content, and improved insulin sensitivity and β-cell function [2]. This initial trial, including a small sample size of 11 subjects, was followed by a more extensive landmark study, the Diabetes Remission Clinical Trial (DiRECT). It was the first trial to show that remission of T2D by VLCDs is realizable in a primary care setting, making it a practical target in treating patients with overweight or obesity who had been diagnosed with T2D within the past 6 years. 46% and 36% remission rates were reported after one and two years, respectively [6, 7]. Comparable approaches were demonstrated to be effective in diverse populations like the Middle East and North Africa, South Asia, Australia, and New Zealand [8–11] and even in non-obese individuals [1]. The remission rates after one year were between 40% – 70% [1, 8–10]. Interestingly, participants who achieved remission of T2D within the very large Look AHEAD trial had improved long-term health outcomes, e.g., considerably lower incidences of chronic kidney and cardiovascular diseases [12] underlining the importance of diabetes remission. However, weight regain is frequent in the long term and the rate of individuals in remission decreased to 13% after five years within the extended intervention of the DiRECT trial [13].

Important factors for the achievement of T2D remission are thought to be substantial weight loss and short duration of diabetes, as individuals that lost the most weight were shown to exhibit the highest chances of remission, and shorter diabetes duration has also been identified to be of importance for the recovery of residual β-cell function [6, 14–16]. For this reason, the above-mentioned T2D remission trials focused on individuals with a diabetes duration not exceeding three or six years [6, 8–10]. Whether such an intervention will demonstrate comparable efficacy in individuals with longer diabetes duration remains therefore understudied. This information is urgently required to develop more tailored interventions. Thus, we aimed to investigate diabetes remission after three months of VCLD in individuals with short (≤4 years) and long (≥8 years) duration of T2D.

Although no macronutrient composition appears to be superior for weight loss in people with T2D [3], lower mean glucose levels and HbA1c were reported after a low-carbohydrate diet induced weight loss compared to a high-carbohydrate diet induced weight loss, despite similar body weight reduction [17]. Nevertheless, the macronutrient profile of VLCD programs for diabetes remission has not received much attention. Therefore, we investigated the impact of two VLCDs differing in their macronutrient composition.

## Research Design and Methods

### Participants

Between September 2020 and November 2022, 52 individuals with overweight or obesity and T2D without insulin treatment were recruited via advertisement in local newspapers, websites, displays on campus, in medical practices in the region, and from the outpatient centers of the Department of Endocrinology at Charité – Universitätsmedizin Berlin, Germany. Individuals were eligible for study participation if they had a BMI above 27 kg/m^2^, were between 18-80 years old, had a diabetes duration since diagnosis of either up to four years or at least eight years (diagnosis based on medical records, or an HbA1c ≥6.5 % (48 mmol/mol), or fasting plasma glucose ≥126 mg/dL without any antidiabetic medication), no prior insulin treatment and were willing to follow a very low-calorie formula diet intervention. Exclusion criteria were as follows: patients without health insurance, patients with type 1 diabetes, insulin treatment, weight loss of more than 5 kg within the last three months before inclusion, alcohol or drug abuse, use of systemic glucocorticoids, pregnancy or breastfeeding, chronic conditions that are incompatible with the experimental design such as known severe hepatic disease or liver cirrhosis, severe psychiatric disease, systemic infection, untreated endocrine disease, severe disease of kidney or heart, advanced diabetic retinopathy, eating disorder, or food allergy to compounds of the formula diets. The *FAsting-induced Immune-metabolic Remission of type 2 diabetes* (FAIR) study protocol was approved by the ethics committee of the Charité-Universitätsmedizin Berlin (EA4/019/20, date of approval: 12.05.2020) and conducted in accordance with the principles of the Declaration of Helsinki 1975, as revised in 2013. Prior to the study all participants gave written informed consent. The FAIR study was registered at ClinicalTrials.gov: NCT05295160.

### Study design

In this randomized controlled trial with parallel group design participants followed a very low-calorie diet (VLCD) for three months and underwent study visits at baseline (V1), performed approximately one week after an initial screening, and after three months of VLCD (V3). At both study visits detailed phenotyping was performed along with medical and nutritional consultation. Antidiabetic medication was discontinued three (oral medication) or seven days (GLP-1 receptor agonist injections) prior to V1 and for the entire three months intervention. During the VLCD, adjustments to antihypertensive drugs, including dose reduction or discontinuation, were made on an individual basis in consultation with the study physician.

After study inclusion, participants were randomly assigned to a meal replacing formula diet, either OPTIFAST^®^ professional (Nestlé Health Science, Frankfurt am Main, Germany) or HEPAFAST^®^ (Nestlé Health Science, division BODYMED AG, Kirkel, Germany). Randomization was performed by the study team using a computer-generated (Microsoft Excel 2019, Microsoft Corporation, Redmond, USA) stratified randomization list. Stratification considered age (<50 years or ≥50 years), sex (female or male), diabetes duration (≤4 years or ≥8 years) and BMI (<30 kg/m^2^ or ≥30 kg/m^2^) at screening.

### Outcomes

Prespecified primary outcome was the remission of T2D, defined as fasting plasma glucose (FPG) <126 mg/dL after three months off all antidiabetic medications, in individuals with a diabetes duration of up to 4 years compared to individuals with a diabetes duration of at least 8 years since diagnosis. In accordance with the consensus report published in 2021 after the start of our trial, favoring an HbA1c <6.5 % without continuation of the antihyperglycemic medication for at least three months as usual defining measurement for diabetes remission [21], we additionally used that definition as a secondary analysis. Secondary outcomes were i) remission rates of both formula diets, ii) additional metabolic and anthropometric parameters predicting diabetes remission.

### Dietary Intervention

The meal replacement diet consisted of three to four formula shakes and a 240 g portion of non-starchy vegetables, resulting in a daily caloric intake of about 600 kcal/d for women and about 800 kcal/d for men. The participants were randomly assigned to consume either OPTIFAST^®^ professional or HEPAFAST^®^ products as meal replacement. Prior to consumption, the OPTIFAST^®^ professional powder was dissolved in water, whereas the HEPAFAST^®^ powder was mixed with a predefined amount of low fat milk (for men: twice a day 300 ml and one time 250 ml, for women: three times 150 ml) or, for variety, with predefined quantities of equivalent products (e.g. low fat joghurt). The resulting macronutrient composition was 25 Energy% (EN%) fat, 35 EN% carbohydrates, 3 EN% fiber, 37 EN% protein in the OPTIFAST^®^ group and 22 EN% fat, 28 EN% carbohydrates, 6 EN% fiber, 44 EN% protein in the HEPAFAST^®^ group. The study center provided the formula products free of charge over the entire study period.

The beverages allowed were unsweetened water, coffee, and tea. No sugar substitutes or alcoholic beverages were allowed during the intervention. The non-starchy vegetables could be consumed raw, cooked, or baked, however preparation with fat or sugar was prohibited. Participants were encouraged to maintain their usual physical activities. Subsequent to the three months of VLCD participants received guidance for a structured food reintroduction phase of two weeks.

### Phenotyping

#### Anthropometry

Anthropometric ascertainment was performed identically at baseline (V1) and after 12-weeks VLCD intervention (V3). Body weight was measured in underwear only with a digital scale (KERN MXS, KERN & SOHN, Balingen-Frommern, Germany) and body height was measured with a stadiometer (Seca, Hamburg, Germany). These measurements were used to calculate the body mass index (BMI), defined as the weight in kilograms divided by the square of the height in meters. Waist and hip circumference were measured with a non-elastic tape two times consecutively and a mean value was calculated, which was used to compute the waist-to-hip ratio (WHR). Body composition was assessed via air displacement plethysmography according to manufacturer instruction (BOD POD, COSMED Srl, Rome, Italy). Participants were measured wearing only underwear and a bathing cap. They were instructed to take off any other clothing and remove jewelry. Blood pressure was measured with a boso ABI-system 100 (BOSCH + SOHN GmbH, Jungingen, Germany).

#### Laboratory analysis

At study visits, fasting blood samples were drawn in the morning (>10 h since last food intake). Determination of routine lab parameters (HbA1c, total cholesterol, HDL cholesterol, LDL cholesterol and triglycerides) was performed at Labor Berlin – Charité Vivantes GmbH, Berlin, Germany. After fasting blood collection, a Mixed Meal Tolerance Test (MMTT) was started by consumption of 243 ml Nestlé Resource^®^ Protein Vanille (Nestlé Health Science, Frankfurt am Main, Germany) mixed with 12 ml sugar solution (Jubin Pharma, Bochum, Germany) resulting in 21 EN% fat, 54 EN% carbohydrate and 25 EN% protein (366 kcal) as standardized meal challenge. Blood was drawn 30, 60, 90, 120 and 180 minutes after consumption. Participants remained resting in semi-supine position during the entire MMTT. Serum samples were kept clotting for 20-30 min at room temperature while plasma samples containing EDTA were centrifuged immediately after collection at 1.800 x g for 10 min at 4 °C and both were stored at -80 °C until analysis. Insulin and C-peptide were measured in plasma samples with specific human ELISA kits (Mercodia, Uppsala, Sweden, Insulin: within assay CV 3.4%, total assay CV 4.5%, C-peptide: within assay CV 3.6%, total assay CV 5.0%). Blood glucose was measured in plasma samples using the hexokinase method (ABX Pentra 400 device (HORIBA Europe, Montpellier, France)).

The homeostasis model assessment-estimated insulin resistance (HOMA-IR) was calculated as fasting insulin [mU/L] multiplied with fasting glucose [mmol/L] divided by 22.5 [18].

### Statistical analysis

Prior to data analysis, a test of plausibility was performed and unusual values that were outside of the 1.5-fold interquartile range and not physiologically plausible were declared as extreme outliers and not considered in further analysis. The Shapiro-Wilk test was used to assess variables for normal distribution. Unless stated otherwise, data are reported as numbers for categorical variables, as mean ± standard deviation (SD) for normally distributed variables and as median (interquartile range) for non-normally distributed variables.

To test for differences in baseline characteristics and changes between diabetes duration, formula diet, and remission groups we used chi-square test for categorical variables, independent samples t-test for normal distributed data and Mann-Whitney U-test for skewed data. In cases of missing data, pairwise deletion was applied. For comparisons between the three weight loss categories (<10 kg, 10-15 kg, `≥`15 kg) we used chi-square test, Kruskal-Wallis-test, or ANOVA with Bonferroni correction, when appropriate. We calculated the area under the curve (AUC) of the MMTT for glucose, insulin, and C-peptide using the trapezoidal method.

To further examine which variables had an impact on T2D remission, binary logistic regression models were used. As the macronutrients show a high multicollinearity with each other, binary logistic regressions were performed for each macronutrient separately. All models were adjusted for sex and age. Despite the macronutrients, identical independent variables were included (baseline C-Peptide, baseline fasting plasma glucose, Δ weight, diabetes duration) in each regression model. Odds ratios with 95% confidence intervals (CI) are described for interpretation of the regression results.

Statistical analyses were processed using SPSS 28.0 (SPSS Inc., Chicago, IL, USA) and p-values < 0.05 were considered significant. The graphs were generated with GraphPad Prism 9 (GraphPad, San Diego, CA, USA).

### Sample size calculation

The sample size was calculated for the primary outcome: remission of T2D compared between individuals with short and long diabetes duration. As diabetes duration is a fixed characteristic of the participants and we aimed for a randomized trial, participants were randomized into two different formula diet groups and the diabetes duration (≤4 years or ≥8 years) was considered for stratification. The initial sample size calculation was based on the results by Steven et. al. [15]. With an expected proportion of diabetes remission three months after starting the fasting program of 87% (diabetes duration ≤4 years) and 50% (diabetes duration ≥8 years), at least 24 patients per group are required to achieve a power of 80% and a significance level of 0.05, using a two-sided chi-square test. A dropout rate of 25% was expected, resulting in an estimated sample size of n=64 (32 participants per group). The sample size calculation was carried out with nQuery 8.4.1.0 (Statsols, Boston, USA).

Due to COVID-19 pandemic related contact restrictions and multiple lockdowns of the outpatient study center at Charité – Universitätsmedizin Berlin between 2020 and 2022, it was not possible to enroll 64 participants within the funded period. Thus, the study was completed with 52 participants enrolled, out of which 47 completed the trial and were included in the present analysis (Figure 1). To evaluate the statistical power of our study with the actual analyzed participant numbers, we performed a post-hoc power analysis [19, 20]. With a proportion of T2D diabetes remission of 82% in the diabetes duration ≤4 years group (33 subjects) and 50% in the diabetes duration ≥8 years group (14 subjects) and a significance level of 0.05, we achieved a power of 61%.

**Figure 1:**
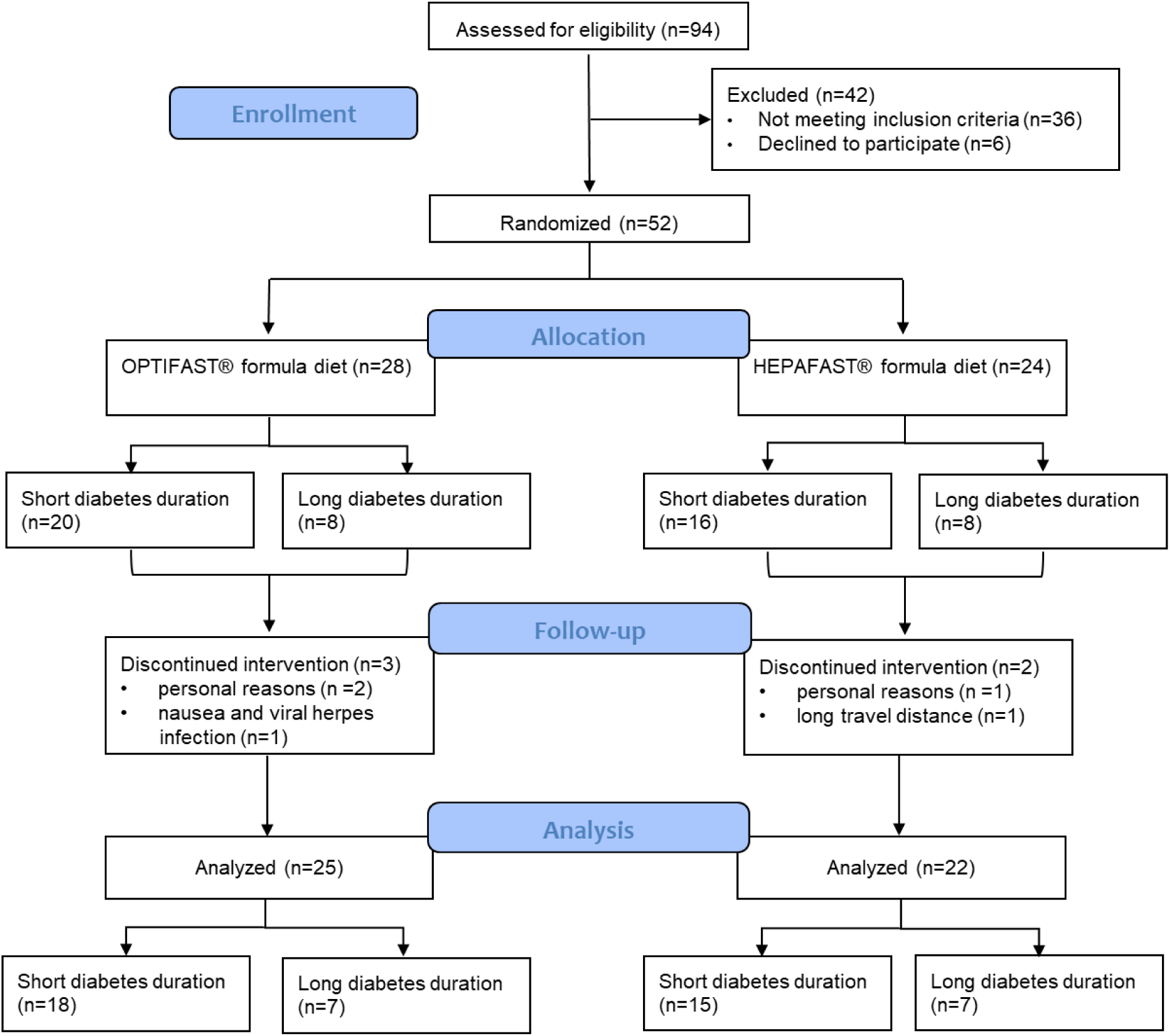
CONSORT Flow chart of the FAIR study.

## Results

### Participant characteristics

Fifty-two subjects with a BMI above 27 kg/m^2^ and non-insulin treated T2D were enrolled in this randomized controlled trial, 28 were randomly allocated to consume OPTIFAST^®^ and 24 to consume HEPAFAST^®^ formula products as meal replacement. Thirty-six of the participants had their T2D diagnosed within the last four years and 16 individuals had it diagnosed at least eight years before study participation. The flow of participants through each stage of the FAIR trial is shown in figure 1. Five participants discontinued the VLCD due to varying reasons (e.g. long travel distance), thus 47 individuals completed the intervention and were analyzed. Their baseline characteristics are shown in table 1 and table S1. In detail participants with a diabetes duration of at least eight years were on average eight years older, had a lower body weight as well as higher FPG and lower C-peptide levels.

**Table 1:** Baseline clinical parameters of all participants and the short (≤4 years) and long (≥8 years) diabetes duration groups.

| Parameter | All participants<br>n = 47 | Short diabetes<br>duration<br>n = 33 | Long diabetes<br>duration<br>n = 14 | p-value |
| --- | --- | --- | --- | --- |
| Sex [female/male] | 25/22 | 15/18 | 10/4 | 0.103 |
| Age [years] | 55 ± 11 | 52 ± 11 | 60 ± 10 | 0.024 |
| Formula diet group<br>[OPTIFAST<sup>®</sup>/HEPAFAST<sup>®</sup>] | 25/22 | 18/15 | 7/7 | 0.775 |
| Diabetes duration [years] | 3(1-8) | 2(1-3) | 10(8-11) | 0.000 |
| Weight [kg] | 106.8 ± 16.8 | 111.6 ± 16.3 | 95.4 ± 11.9 | 0.002 |
| BMI [kg/m<sup>2</sup>] | 35.8 ± 5.3 | 36.8 ± 5.7 | 33.5 ± 3.5 | 0.050 |
| Fat mass [%] | 43.6(36.7-51.1) | 44.3(36.0-51.2) | 43.0(38.3-51.2) | 0.857 |
| Fat free mass [%] | 56.4(48.9-63.3) | 55.7(48.8-64.0) | 57.0(48.8-62.7) | 0.857 |
| Waist circumference [cm] | 117 ± 13 | 120 ± 13 | 112 ± 9 | 0.063 |
| WHR | 0.98 ± 0.08 | 0.99 ± 0.08 | 0.97 ± 0.09 | 0.468 |
| Systolic BP left arm [mmHg] | 137(127-151) | 142(126-154) | 137(133-148) | 0.576 |
| Diastolic BP left arm [mmHg] | 93 ± 10 | 93 ± 11 | 92 ± 8 | 0.302 |
| Fasting insulin [mU/L] | 14.3(8.2-19.1) | 15.4(9.3-21.7) | 9.8(5.4-15.7) | 0.073 |
| Fasting glucose [mg/dL] | 150.6(139.3-183.8) | 147.9(132.4-163.2) | 174.5(148.2-239.4) | 0.027 |
| Fasting C-peptide [pmol/L] | 1127 ± 477 | 1338 ± 486 | 962 ± 338 | 0.015 |
| HbA1c [%] | 6.8(6.2-7.5) | 6.6(6.1-7.3) | 7.1(6.7-8.4) | 0.056 |
| HbA1c [mmol/mol] | 50.3(40.0-57.9) | 48.2(43.0-55.5) | 53.9(49.0-67.9) | 0.061 |
| HOMA-IR | 5.66(3.34-8.89) | 5.68(3.46-8.61) | 4.01(1.93-9.82) | 0.515 |
| Fasting total cholesterol<br>[mg/dL] | 192 ± 40 | 195 ± 32 | 185 ± 53 | 0.509 |
| Fasting HDL cholesterol<br>[mg/dL] | 47(40-53) | 43(38-52) | 52(46-68) | 0.029 |
| Fasting LDL cholesterol<br>[mg/dL] | 119 ± 36 | 121 ± 29 | 113 ± 48 | 0.563 |
| Fasting triglycerides [mg/dL] | 151(106-219) | 155(107-237) | 150(96-181) | 0.312 |

Values are shown as mean ± SD for normal distributed variables and as median (interquartile range) for skewed variables. P-values shown for between-group difference. BMI, Body Mass Index; BP, Blood pressure; HOMA-IR, Homeostasis Model Assessment-Insulin Resistance; WHR, Waist-to-hip-ratio.

Thirty-five subjects were treated with antidiabetic medications prior to their study participation: 15 patients were on monotherapy (ten with metformin, two with GLP-1 receptor agonists, one with SGLT2 inhibitor, two with DPP4 inhibitors), 20 patients were on combination therapy (seven with metformin + GLP-1 receptor agonists, five with metformin + DPP4 inhibitors, one with SGLT2 inhibitor + DPP4 inhibitor, four with metformin + SLGT2 inhibitors, one with metformin + sulfonylurea, one with metformin + GLP-1 receptor agonist + SGLT2 inhibitor, one with metformin + DPP4 inhibitor + SGLT2 inhibitor). Previous GLP-1 receptor agonist treatment was equally distributed between the two formula diet groups.

### Changes in body weight and composition

On average study participants reduced their body weight by 15.6 ± 5.2 kg. Twenty-five subjects lost more than 15 kg, thirteen lost between 10 – 15 kg and nine subjects lost less than 10 kg. The mean weight loss and the proportional distribution within the three weight loss categories (Figure 2 and Figure 3) was comparable between the short and long diabetes duration group (short: -15.2 ± 5.8 kg vs long: -13.9 ± 4.8 kg, p = 0.473) and between the two formula diet groups (OPTIFAST®: -15.3 ± 5.7 kg vs HEPAFAST®: -14.2 ± 5.3 kg, p = 0.477).

**Figure 2:**
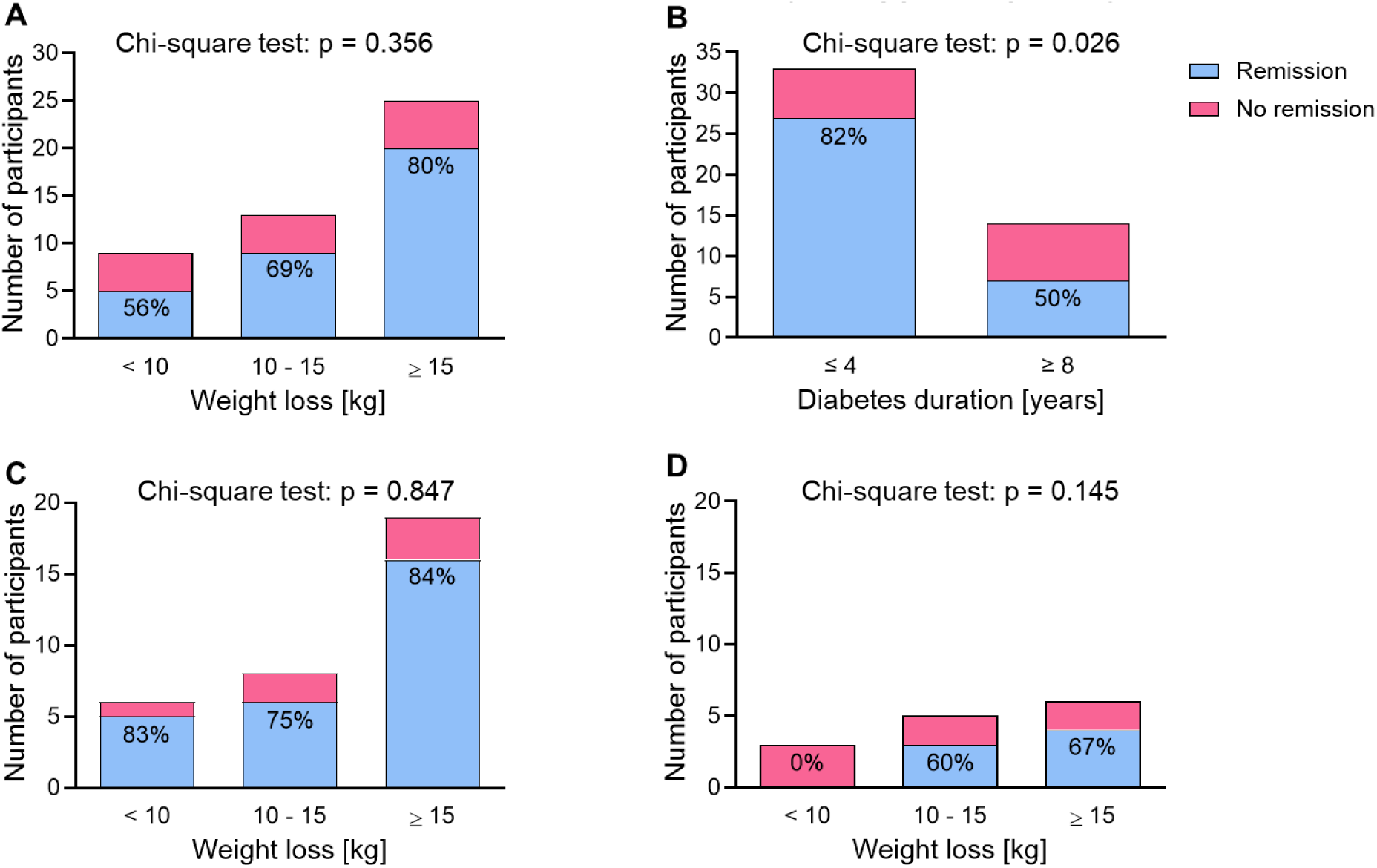
Remission of T2D in response to three months of VLCD. **A:** Total cohort. T2D remission was closely related to the degree of weight loss. **B:** Remission of T2D compared between the short and long diabetes duration group. **C:** T2D remission rates in relation to weight reduction in the short diabetes duration group (≤4 years). **D:** T2D remission rates in relation to weight reduction in the long diabetes duration group (≥8 years).

**Figure 3:**
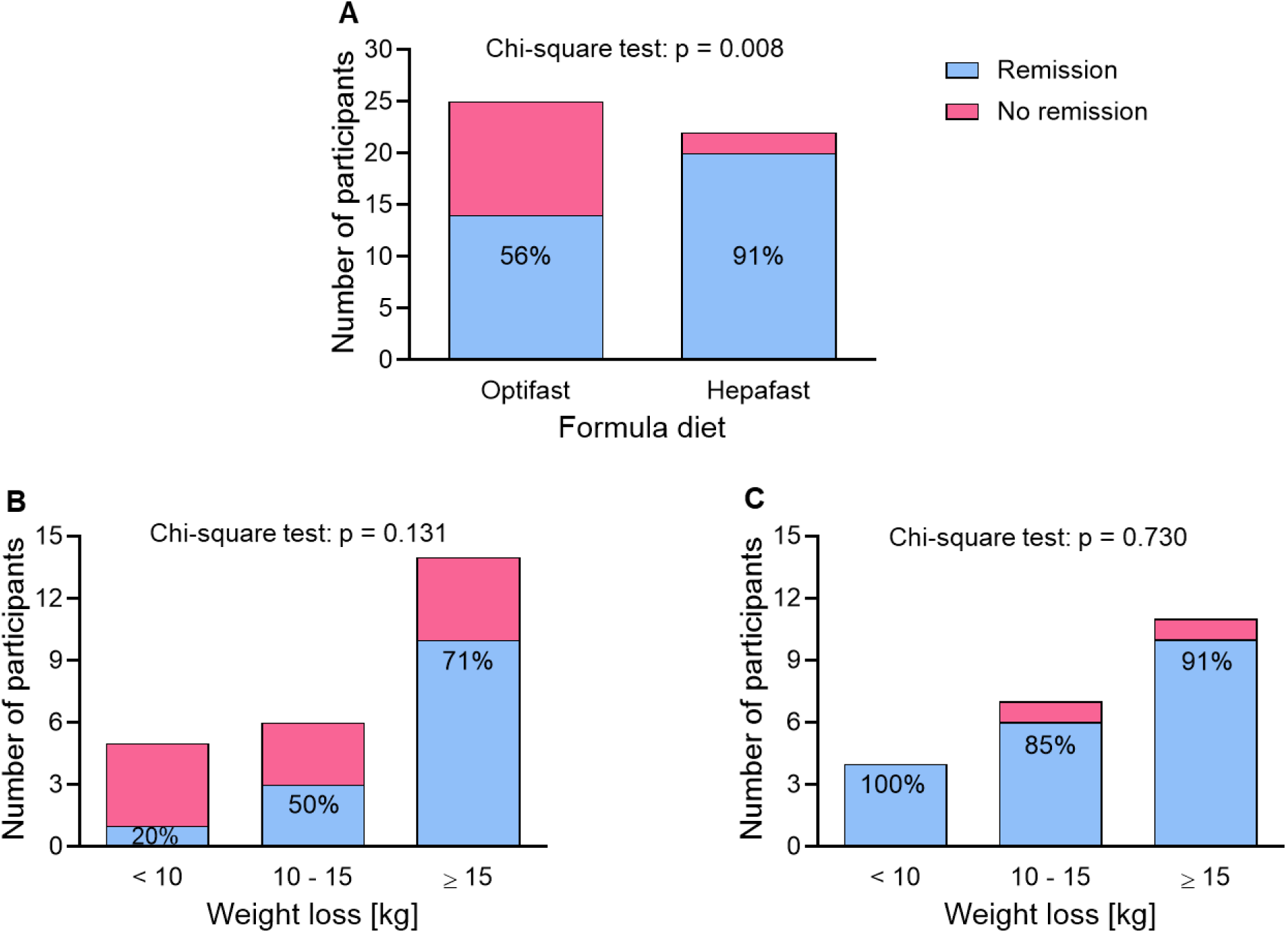
Remission of T2D in the formula diet groups. **A:** Remission of T2D compared between the OPTIFAST^®^ and HEPAFAST^®^ group. **B:** T2D remission rates in relation to weight reduction in the OPTIFAST^®^ group. **C:** T2D remission rates in relation to weight reduction in the HEPAFAST^®^ group.

The participants’ weight loss was mainly due to a reduction in fat mass (-13.6 ± 4.0 kg) over the three months of VLCD. Nonetheless, the participants also lost 2.1 ± 2.9 kg of their fat-free mass. There were no differences between the two diabetes duration groups (p=0.698) or between the two diet groups (p=0.627) in terms of reduction in fat and fat free mass in response to the intervention (data not shown).

### Remission of type 2 diabetes

Thirty-four out of the 47 study completers achieved a remission of T2D, defined as fasting glucose levels below 126 mg/dL at the end of the intervention. This results in a 72% remission rate in the total cohort. We analyzed the remission rates for each weight loss category and found that the remission of T2D was slightly related to the degree of weight loss, even though the differences between the three weight loss categories were not significant (p=0.356), which might be caused by the relatively small sample size (Figure 2 A).

Using the definition of diabetes remission published in 2021, which favors an HbA1c < 6.5 % without continuation of the usual antihyperglycemic medication for at least three months as the main defining measurement [21], we found a remission rate of 91 % (Figure S1).

We found a significantly higher remission rate of T2D in subjects with a diabetes duration of 4 years or less in comparison with subjects that had their T2D diagnosed for at least 8 years (Figure 2B). Especially in individuals with longer diabetes duration a weight loss of more than 10 kg appears to be essential to achieve a remission of T2D (Figure 2C and D).

### Impact of the type of formula diet on the remission of type 2 diabetes

As the study participants consumed either OPTIFAST^®^ or HEPAFAST^®^ formula as meal replacement, we investigated whether the T2D remission rates differed between the two formula diet groups. Surprisingly, we found a significantly higher remission rate of T2D in the HEPAFAST^®^ group (p=0.008) (Figure 3A). In the OPTIFAST^®^ group the remission rates increased progressively with higher weight loss after three months, whereas this trend was not found in the HEPAFAST^®^ group (Figure 3B and C).

### Impact of further parameters on the remission of type 2 diabetes

We then compared baseline parameters (Table 2) as well as the changes of these parameters over the three months of VLCD (Table S2) between the study participants that achieved a T2D remission and those who did not. Individuals that were in remission at the end of the intervention had significantly lower fasting glucose levels at baseline. In addition, they had significantly higher fasting C-peptide levels and a greater C-peptide area under the curve (AUC) (Table 2). When comparing changes of the parameters we found a significantly stronger decrease of fasting triglycerides (-78[- 114 - -29] mg/dL vs. -26[-48 – 0] mg/dL, p=0.027) and insulin AUC in the remission group (-2934 ± 672 mU/L x 180 min vs. -1010 ± 495 mU/L x 180 min, p=0.026). We found no statistically significant difference between the remission and non-remission group in the FPG improvement (-39.9[-58.4 - -19.7] mg/dL vs. -22.5[-86.1 - 0.5] mg/dL, p=0.348) (Table S2).

**Table 2:** Baseline characteristics of individuals with T2D remission and those with no T2D remission at the end of the three months intervention period.

| Parameter | Remission<br>n = 34 | No remission<br>n = 13 | p-value |
| --- | --- | --- | --- |
| Sex [female/male] | 16/18 | 9/4 | 0.173 |
| Age [y] | 55(48-61) | 58(47-66) | 0.528 |
| Weight [kg] | 106.4(98.9-117.7) | 96.0(88.6-109.8) | 0.057 |
| BMI [kg/m<sup>2</sup>] | 36.4 ± 5.3 | 34.3 ± 5.2 | 0.216 |
| Fat mass [%] | 41.9(36.7-51.9) | 45.8(34.8-49.65) | 0.762 |
| Fat free mass [%] | 58.1(48.1-63.3) | 54.2(50.4-65.2) | 0.762 |
| Waist circumference [cm] | 117(110-124) | 110(106-123) | 0.221 |
| WHR | 0.99 ± 0.08 | 0.98 ± 0.09 | 0.707 |
| Systolic BP left arm [mmHg] | 139 ± 14 | 145 ± 21 | 0.275 |
| Diastolic BP left arm [mmHg] | 93 ± 10 | 92 ± 11 | 0.867 |
| Fasting insulin [mU/L] | 14.9(9.4-20.6) | 9.8(5.9-16.8) | 0.111 |
| Fasting glucose [mg/dL] | 149.5(132.7-165.4) | 187.7(144.6-251.4) | 0.017 |
| Fasting C-peptide [pmol/L] | 1314 ± 513 | 997 ± 257 | 0.005 |
| HbA1c [%] | 6.6(6.1-7.3) | 6.9(6.6-9.1) | 0.082 |
| HbA1c [mmol/mol] | 48.4(43.5-56.2) | 52.0(49.0-75.1) | 0.074 |
| HOMA-IR | 5.67(3.64-7.71) | 4.19(2.15-9.74) | 0.755 |
| AUC glucose [mg/dL x 180 min] | 33531(28919-37359) | 37973(30569-52678) | 0.067 |
| AUC insulin [mU/L x 180 min] | 8666(6191-16400) | 7606(4291-9988) | 0.087 |
| AUC C-peptide [nmol/L x 180 min] | 392(309-533) | 321(234-345) | 0.035 |
| Fasting TC [mg/dL] | 195 ± 35 | 183 ± 50 | 0.338 |
| Fasting HDL-C [mg/dL] | 44(40-52) | 50(40-62) | 0.258 |
| Fasting LDL-C [mg/dL] | 120 ± 32 | 114 ± 45 | 0.575 |
| Fasting TG [mg/dL] | 159(104-245) | 149(100-173) | 0.225 |

Values are shown as numbers, mean ± SD for normal distributed variables and as median (interquartile range) for skewed data. AUC, area under the curve; BMI, Body Mass Index; BP, Blood pressure; HDL-C, High-density lipoprotein cholesterol; HOMA-IR, Homeostasis Model Assessment-Insulin Resistance; LDL-C, Low-density lipoprotein cholesterol; TC, Total cholesterol; TG, Triglycerides; WHR, Waist-to-hip-ratio.

### Predicted probability of type 2 diabetes remission

Finally, we aimed to analyze the impact of different markers on diabetes remission via binary logistic regression models. This was adjusted for possible confounders as sex and age. In addition to lower FPG and higher C-peptide levels, the formula diet composition, in detail higher protein and fiber content together with less fat and carbohydrates, could be identified as potential predictors of T2D remission (Figure 4).

**Figure 4:**
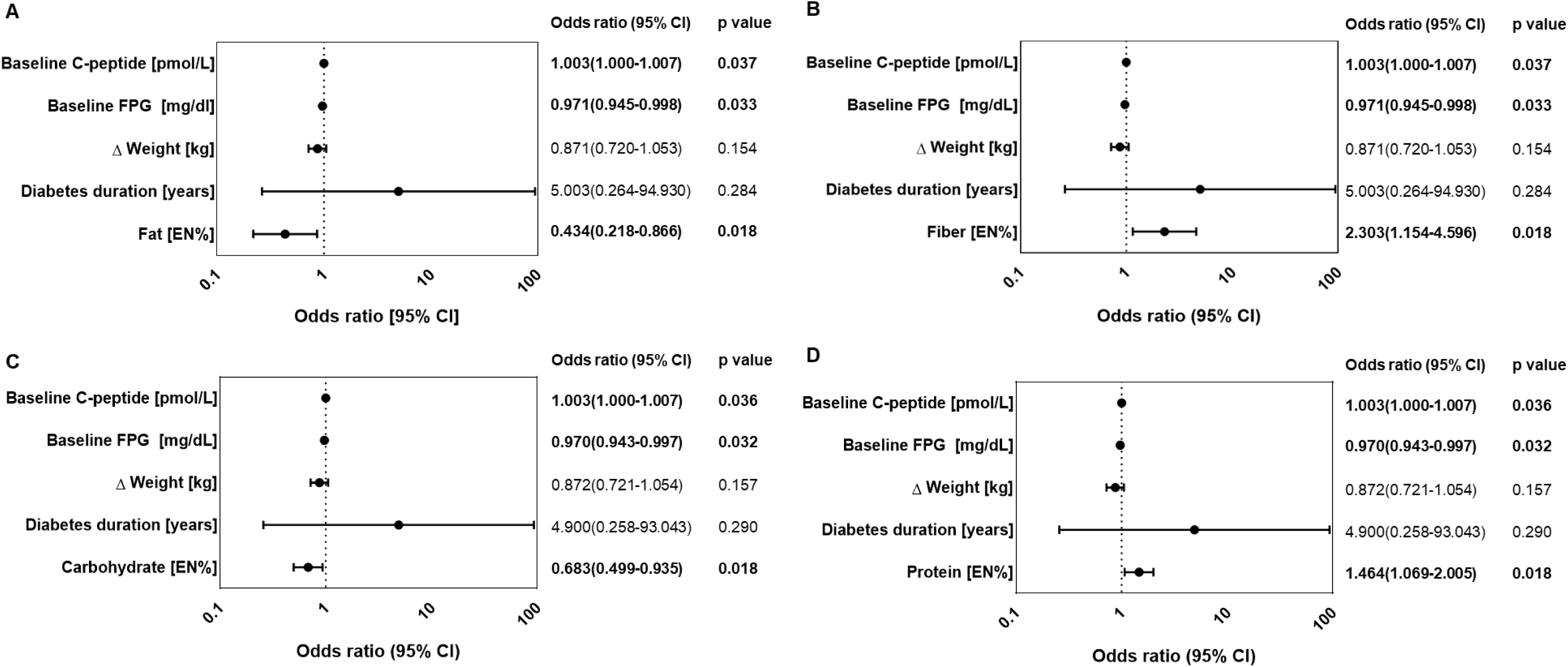
Predicted probability for remission of T2D. Binary logistic regression was applied for each macronutrient separately. All odels were adjusted for age and sex. **A:** Fat intake [EN%]. **B:** Fiber intake [EN%]. **C:** Carbohydrate intake [EN%]. **D:** Protein intake EN%].

## Discussion

Our study demonstrates that a very low-calorie formula diet for three months, combined with discontinuation of all antidiabetic medications, is sufficient to induce significant diabetes remission in nearly three-quarters of participants with T2D and overweight or obesity. Although the effect is somewhat reduced, it can be also observed in subjects with a longer diabetes duration. Diabetes remission goes along with a substantial weight loss induced by VLCD intervention. The remission rate observed in the present trial is slightly higher compared to previous one-year trials, including the DiRECT trial [6, 8–10]. This is most likely due to a different timing of diabetes remission ascertainment. It is known that weight regain is a common problem following VLCD interventions and is associated with diabetes relapse [7, 22]. In consequence, more subjects are expected to be in remission directly after the VLCD phase than after a subsequent food reintroduction and weight maintenance phase, as reported in the DiRECT-Australia trial [9].

Another aspect to consider is the usage of different criteria to assess remission of T2D. Discontinuation of antidiabetic medication is a prerequisite in almost all trials. In our trial, diabetes remission was predefined as FPG below 126 mg/dL, as the consensus report favoring HbA1c as the main defining criterion was published after the start of our study, whereas the other studies considered participants with an HbA1c below 6.5% in remission. When applying the HbA1c cut-off, the remission rate in the current trial is higher and very close to the rate reported after 12 weeks in the DiRECT-Australia study [9].

Most importantly, prior T2D remission trials included individuals with a short diabetes duration only and excluded individuals with a diabetes duration greater than three or six years [1, 6, 8–10]. In contrast, we included also subjects with a longer diabetes duration. Thereby we could demonstrate a substantial remission rate also in this cohort, even it is somewhat smaller. Solely one previous study with an 8-week VLCD intervention included patients with T2D of short (<4 years) and long duration (>8 years). The findings of this trial, demonstrating that 87% of individuals with short diabetes duration and 50% with long diabetes duration achieved FPG levels below 126 mg/dL, are in close accordance with our study results [15]. The average weight loss was similar between the short and long diabetes duration groups in both studies. Another similarity between the two trials is that participants with short diabetes duration were significantly younger and had lower baseline FPG levels [15]. Taken together, current data indicates that a shorter diabetes duration appears to enhance the chances of T2D remission through a VLCD intervention. Nevertheless, it remains a promising approach for approximately half of those with longer diabetes duration.

Given the impact of different macronutrient patterns on glucose metabolism [4, 17], we aimed to disentangle the impact of those macronutrients in the context of a VLCD approach. To our knowledge this is the first study to directly compare two liquid formula diets with different macronutrient profiles consumed as total meal replacement to achieve diabetes remission. Remarkably, a significant difference in the remission rates was found between the two formula diet groups. The group that consumed a liquid meal replacement with moderately higher carbohydrate and fat and, therefore, slightly less protein and fiber content had, despite similar weight loss, a significantly lower remission rate. In accordance, a low-carbohydrate weight loss intervention exhibited lower mean glucose levels and HbA1c compared to weight loss induced by a high-carbohydrate diet after six months and one year [17]. Moreover, higher intakes of fiber were shown to improve measures of glycemic control in individuals with diabetes [4, 23]. In addition, a higher protein intake during weight loss interventions may provide moderate advantages compared to low protein intake in people with T2D [24]. Our data gives the first indication that macronutrient profiles are important in VLCD regimes and could be used to optimize remission success. Given the limited sample size of our trial, further and larger trials are clearly required to verify our findings.

Finally, we aimed to identify individual characteristics responsible for and predicting diabetes remission. As seen in a previous trial [25], lower baseline FPG was identified as an important factor for achieving diabetes remission after three months. However, other factors also emerged as important for diabetes remission in the present study. C-peptide levels were significantly higher prior to the intervention in individuals who achieved a remission. Moreover, the postprandial rise in insulin was reduced stronger in participants who achieved T2D remission. The stronger decline in insulin excursion was also observed in earlier studies and attributed to higher baseline insulin levels [25, 26] which was also present in our cohort but not significant. Higher C-peptide levels at baseline in participants with diabetes remission may be interpreted as greater secretory β-cell capacity, as prior studies showed that a recovery of the insulin secretory capacity defined individuals that returned to normoglycemia [25, 26]. Although HOMA-IR did not differ between participants with and without T2D remission, the stronger decline in triglycerides observed in individuals with diabetes remission is frequently considered as a sign of improved insulin sensitivity [27, 28]. However, we did not use sophisticated methods to differentiate the effects on organ specific insulin sensitivity or insulin secretion. Thus, further trials are required to deeper characterize the changes of insulin secretion and sensitivity upon remission. Meanwhile, the higher C-peptide levels observed in patients achieving T2D remission pointed out, that a substantial secretory function is required to achieve diabetes remission, even in subjects with longer diabetes duration.

In contrast to the present study, a previous regression from the DiRECT trial identified weight loss as the most important predictor for the remission of T2D and fasting C-peptide did not predict remission success [16]. Weight loss was not found to be a potential predictor of T2D remission in the present FAIR study which may be caused by the smaller sample size and the fact that the participants already had lower Hb1Ac and FPG levels at baseline, indicating less severe diabetes cases. In addition, the FAIR study participants achieved greater weight loss than those in the DiRECT trial, which minimizes the modulating effect of weight loss, too. Nevertheless, sufficient weight loss was also important for diabetes remission in the present trial. Even though diabetes duration is an important factor for the remission of T2D and has been shown to inversely correlate with β-cell capacity, diabetes duration did not emerge as a predictor of remission, neither in this nor in the DiRECT study with a smaller spectrum of diabetes duration [16, 22]. Based on this, one could argue that the impact of diabetes duration on remission success is rather secondary and mediated via higher age, higher FPG and HbA1c, and lower C-peptide levels present in individuals with longer diabetes duration. Our results underline that diabetes remission remains a reachable target even in people with a long duration of diabetes, and future studies should further explore the role of diabetes duration in the remission of T2D.

The study findings should be interpreted considering several limitations. First, the study was conducted during the COVID-19 pandemic, and contact restrictions and multiple lockdowns of our endocrinology outpatient study center at Charité – Universitätsmedizin Berlin made it impossible to enroll the initially estimated sample size of 64 participants within the funded period. Moreover, not enough participants with long diabetes duration without insulin treatment were recruited, resulting in unmatching sizes of the diabetes duration groups. A post-hoc power analysis [19, 20] considering the smaller and unequal group sizes resulted in a power of 61% for the comparison between the diabetes duration groups. Second, baseline fasting plasma glucose levels significantly differed among the two diabetes duration groups. Further matching based on glycemic control or diabetes treatment would have created an unrepresentative group defined by diabetes duration [15]. Third, this study deliberately did not include a control group to not unnecessarily increase the required participant number for ethical reasons. A control group may no longer be required, given the prior evidence on the effectiveness of intensive VLCDs in achieving diabetes remission [6, 8, 9]. Fourth, the study mainly included individuals with Caucasian ethnicity, limiting the generalizability to other populations. However, previous studies in varying populations suggest that similar T2D remission rates can be achieved in populations in the Middle East and North Africa, South Asia, Australia, and New Zealand [8–11]. Finally, minor protocol violations could not be ruled out despite close supervision, as no independent biomarker was available to control for compliance. However, consultation protocols, the high remission rates and weight loss indicate good adherence to the VLCD.

## Conclusion

We show that very low-calorie dietary interventions resulting in significant weight reduction are a very effective approach to achieve the remission of T2D, especially in individuals with low fasting plasma glucose levels, high C-peptide levels, and a more recent diabetes diagnosis. Our results indicate that the macronutrient profile might enhance glucose metabolism improvements even under very low-caloric conditions, but further studies are needed to confirm this finding. Future studies should also address potential differences in long-term responses to define interventions that can effectively support individuals to keep their weight stable and remain in remission over a more extended period.

## Data Availability

All data produced in the present study are available upon reasonable request to the authors.

