## Supplementary material for "Remission of type 2 diabetes can be achieved in short and long-lasting type 2 diabetes: results of a randomized controlled trial in individuals with overweight and obesity"

**Table S1:** Baseline clinical parameters of all subjects and both formula diet groups.

| Parameter | All participants  n=47 | OPTIFAST group  n=25 | HEPAFAST group  n=22 | p-value |
| --- | --- | --- | --- | --- |
| Sex [female/male] | 25/22 | 13/12 | 12/10 | 0.861 |
| Age [years] | 56(49-65) | 60(49-67) | 55(48-59) | 0.084 |
| Diabetes duration [≤4/ ≥8 years] | 33/14 | 18/7 | 15/7 | 0.775 |
| Diabetes duration [years] | 3(1-8) | 3(2-9) | 3(1-8) | 0.390 |
| Weight [kg] | 106.8 ± 16.8 | 108.5 ± 19.8 | 104.8 ± 12.7 | 0.444 |
| BMI [kg/m²] | 35.5(31.7-38.2) | 34.6(31.4-38.8) | 36.9(32.2-38.2) | 0.565 |
| Fat mass [%] | 43.6(36.7-51.1) | 44.3(32.9-52.9) | 43.0(38.4-51.0) | 0.481 |
| Fat free mass [%] | 56.4(48.9-63.3) | 55.7(47.1-67.1) | 57.0(49.0-61.7) | 0.481 |
| Waist circumference [cm] | 117 ± 13 | 119 ± 15 | 115 ± 10 | 0.259 |
| WHR | 0.98 ± 0.08 | 0.99 ± 0.08 | 0.98 ± 0.09 | 0.573 |
| Systolic BP [mmHg] | 140 ± 16 | 141 ± 18 | 140 ± 14 | 0.717 |
| Diastolic BP [mmHg] | 93 ± 10 | 93 ± 12 | 93 ± 9 | 0.870 |
| Fasting glucose [mg/dL] | 150.6(139.3-183.8) | 155.9(140.9-197.8) | 149.6(133.9-168.2) | 0.388 |
| Fasting insulin [mU/L] | 14.3(8.2-19.1) | 12.3(7.0-18.7) | 14.8(9.6-20.7) | 0.359 |
| Fasting C-peptide [pmol/L] | 1125(846-1538) | 1233(905-1416) | 1123(779-1582) | 0.769 |
| HbA1c [%] | 6.8(6.2-7.5) | 6.8(6.5-8.0) | 6.55(6.0-7.4) | 0.254 |
| HbA1c [mmol/mol] | 50.3(40.0-57.9) | 50.7(46.9-63.1) | 47.7(42.4-56.7) | 0.220 |
| HOMA-IR | 5.66(3.34-8.89) | 5.66(2.48-9.35) | 5.71(3.66-8.88) | 0.609 |
| Fasting TC [mg/dL] | 192 ± 40 | 191 ± 43 | 193 ± 37 | 0.909 |
| Fasting HDL-C [mg/dL] | 47(40-53) | 43(40-53) | 48(42-55) | 0.543 |
| Fasting LDL-C [mg/dL] | 119 ± 36 | 118 ± 41 | 119 ± 28 | 0.949 |
| Fasting TG [mg/dL] | 151(106-219) | 155(107-201) | 132(92-234) | 0.654 |

Values are shown as numbers, mean ± SD for normal distributed variables and as median (interquartile range) for skewed variables. P-values for the between-group difference. BP, Blood pressure; BMI, Body Mass Index; HDL-C, High-density lipoprotein cholesterol; HOMA-IR, Homeostasis Model Assessment-Insulin Resistance; LDL-C, Low-density lipoprotein cholesterol; TC, Total cholesterol; TG, Triglycerides; WHR, Waist-to-hip-ratio.

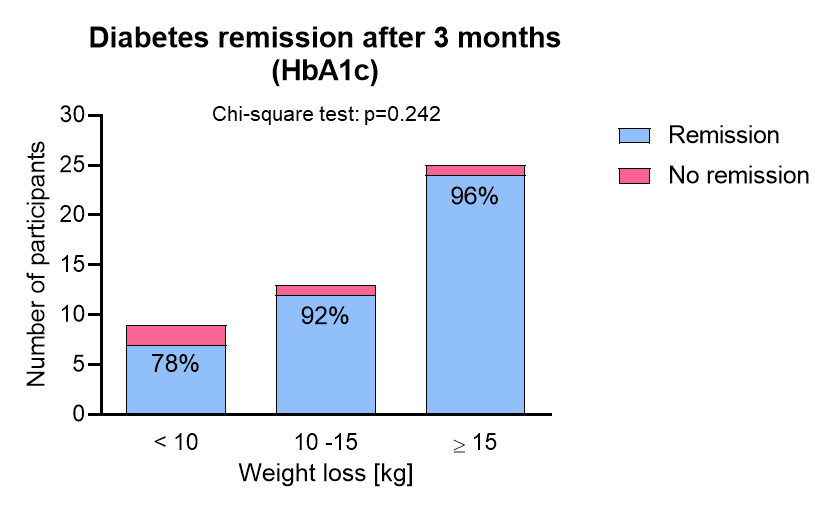

**Figure S1: Remission of T2D defined as HbA1c below 6.5% after three months of VLCD and off all antihyperglycemic medications.** 43 out of the 47 study completers achieved a remission of T2D, resulting in a 91 % remission rate in the total cohort. Similar to the fasting plasma glucose remission definition, the T2D remission defined by HbA1C increased with higher weight loss. However, no significant differences in remission success were found between the weight loss categories (p=0.242).

**Table S2:** Effect of 3 months of VLCD on metabolic and anthropometric markers in subjects with and without T2D remission.

| Parameter Delta V1-V3 | Remission group  n = 34 | No remission group  n = 13 | p-value |
| --- | --- | --- | --- |
| Weight [kg] | -15.4 ± 1.0 | -13.1 ± 1.4 | 0.094 |
| BMI [kg/m²] | -5.6(-6.4 - -4.1) | -4.2(-6.0 - -3.2) | 0.258 |
| Waist circumference [cm] | -13 ± 1 | -13 ± 2 | 0.782 |
| Fat mass [kg] | -14.1 ± 0.6 | -12.2 ± 1.5 | 0.176 |
| Fat free mass [kg] | -2.7(-3.5 - -0.2) | -1.1(-5.1 - -0.8) | 0.889 |
| Systolic BP left arm [mm Hg] | -12 ± 2 | -5 ± 4 | 0.085 |
| Diastolic BP left arm [mm Hg] | -9 ± 1 | -9 ± 3 | 0.916 |
| Fasting glucose [mg/dL] | -39.9(-58.4 - -19.7) | -22.5(-86.1 - 0.5) | 0.348 |
| Fasting insulin [mU/L] | -6.0(-12.2 - -2.4) | -2.8(-6.7 - -1.0) | 0.140 |
| Fasting C-peptide [pmol/L] | -380(-626 - -210) | -235(-421 - -120) | 0.072 |
| HbA1c [%] | -0.8(-1.3 - -0.5) | -0.6(-1.5 - -0.4) | 0.536 |
| HbA1c [mmol/mol] | -8.5(-13.9 - -5.9) | -6.5(-15.5 - -4.4) | 0.542 |
| HOMA-IR | -3.31(-4.88 - -1.84) | -2.08(-5.71- -0.19) | 0.348 |
| AUC glucose [mg/dL x 180 min] | -8438(-12114 - -4174) | -5154(-17999 - -1738) | 0.409 |
| **AUC insulin [mU/L x 180 min]** | **-2934 ± 672** | **-1010 ± 495** | **0.026** |
| AUC C-peptide [nmol/L x 180 min] | -43(-130 - -4) | -14(-59 - 22) | 0.158 |
| Fasting TC [mg/dL] | -32 ± 5 | -21 ± 7 | 0.254 |
| Fasting HDL-C [mg/dL] | -3(-6 - -3) | -5(-15 - 2) | 0.372 |
| Fasting LDL-C [mg/dL] | -21 ± 5 | -14 ± 9 | 0.491 |
| **Fasting TG [mg/dL]** | **-78(-114 - -29)** | **-26(-48 - 0)** | **0.027** |

Values are shown as numbers, mean ± SD for normal distributed variables and as median (interquartile range) for skewed variables. AUC, area under the curve; BMI, Body Mass Index; BP, Blood pressure; HDL-C, High-density lipoprotein cholesterol; HOMA-IR, Homeostasis Model Assessment-Insulin Resistance; LDL-C, Low-density lipoprotein cholesterol; TC, Total cholesterol; TG, Triglycerides; WHR, Waist-to-hip-ratio.
